# Improved diagnosis of rare disease patients through application of constrained coding region annotation and de novo status

**DOI:** 10.1101/2022.08.19.22278944

**Authors:** Hywel J Williams, Chris Odhams, Genomics England

## Abstract

Identifying the pathogenic variant in a rare disease (RD) patient is the first step in ending their diagnostic odyssey. De novo (Dn) variants affecting protein-coding DNA are a well-established cause of Mendelian disorders in RD patients. Constrained coding regions (CCRs) are specific segments of coding DNA which are devoid of functional variants in healthy individuals. Furthermore, the most constrained regions, those in percentile bin >95 (CCR95), are significantly enriched for functional pathogenic variants and could therefore be useful for clinical variant prioritisation.

We aimed to evaluate the diagnostic utility of incorporating Dn, CCR95 and Dn_CCR95 status into the variant prioritisation cascade for RD patients that have undergone genomic sequencing. Using data from the Genomics England 100,000 Genomes Project v12, we selected 3,090 trios that have undergone diagnostic evaluation and been analysed with an advanced Dn identification pipeline. For this analysis we have excluded all non-autosomal variants.

Our analysis shows the diagnostic rate increased from 71% in the full cohort to 81% when evaluating just the CCR95 variants, 84% for Dn variants and 87% for Dn CCR95 variants. Of note, manual evaluation of the Dn CCR95 variants from the undiagnosed patients revealed a putative diagnosis in 69% of patients (27 of 39), with clinical follow up resulting in a diagnosis for a further 11 patients. This takes the overall diagnostic rate for Dn CCR95 variants 90% and suggests application of this metric can prioritise diagnostic variants in undiagnosed patients.

We also identify a striking enrichment of signal in patients with a phenotype of neurology and neurodevelopmental disorders, whereby their diagnostic rate increases from 60% in the whole cohort to 71%, 73% and 74% in the Dn, CCR95 and Dn CCR95 categories respectively.

In summary, we demonstrate the potential clinical utility of performing bespoke Dn analyses of RD patients and for incorporating CCR information into the filtering cascade to prioritise pathogenic variants. We believe such a strategy will aid the identification of pathogenic variants and decrease the time taken to make a diagnosis, thus increasing the overall diagnostic rate by allowing more samples to be analysed over the same time period.

## Introduction

The use of next generation sequencing (NGS) techniques such as gene panels, exome and whole genome sequencing for the diagnosis of patients with rare diseases (RDs) is becoming routine practice in genetic diagnostic laboratories world-wide. This has led to improvements in both the time taken to reach a diagnosis and the overall diagnostic rate (1-4). With the cost of sequencing continuing to fall and automation increasing the number of samples being sequenced, we have reached a position where the interpretation of the data being generated is becoming a bottleneck (5, 6).

The difficulty in interpreting NGS data is down to the sheer volume of variants identified when we compare a person’s DNA to the reference genome and the observation that even though a great number of these variants have characteristics overlapping with pathogenic variants, most, if not all, will be benign (7, 8). In order to simplify the interpretation of NGS data, scientists rely on a suite of annotation tools to help filter out the benign variants and leave a small list of potentially pathogenic variants that can be studied in greater detail. There are now a range of prioritisation tools available to perform this task (reviewed in (7)) and a number of commercial software programs that can semi-automate this process, for example: Qiagen Clinical Insights (https://digitalinsights.qiagen.com/) and Congenica (https://www.congenica.com/).

Even with these prioritisation tools, the task of distinguishing pathogenic from benign variants is time consuming and novel annotation methods are required to improve this efficiency. To this end, Havrilla et al., published a novel resource based on creating a map of constrained coding regions (CCRs) in the human genome (9). These are regions of the coding genome devoid of functional genetic variants. In brief, to determine these regions, the authors utilised the Genome Aggregation Database (gmomAD (10)) resource and invoked the principles of survival bias which, in this context, involved identifying regions of coding sequence that were devoid of functional variants (potentially protein altering) above the average 7bp in size. They next modelled these regions using metrics such as the expected mutation rate based on DNA context and then ranked all the regions into percentile bins. Their analysis of these bins showed that those above the 95^th^ percentile were significantly enriched for known pathogenic variants in ClinVar (11) and mutations underlying developmental disorders. The nature of CCRs is such that because they rely on the appearance of just a single allele, they are ideally suited for augmenting current variant prioritisation methods when evaluating De novo (Dn) variants in studies of autosomal dominant diseases.

Dn variants are a rich source of pathogenicity in RD patients. To date, the majority of large cohort studies have focused on the impact of Dn variants in RD patients with a neurodevelopmental-associated phenotype and have shown unequivocally that they are pathogenic in excess of 50% of patients (12-18). Similar studies in non-neurodevelopmental-associated RD patients have shown Dn variants have a substantial impact but at a lower level, for example ∼8% in patients with congenital heart disease (19). To understand the impact of pathogenic Dn variants in RD patients from across the full phenotypic spectrum requires a large, systematically ascertained and analysed cohort.

To evaluate the utility of applying CCR information in a RD diagnostic setting, we have leveraged the power of the Genomics England (GeL), 100,000 Genomes Project (100KGP) (20). The 100KGP is a landmark genomics project based in the UK that is aligned with the National Health Service (NHS) (see reference for details). The RD component covers the full spectrum of RDs and is therefore well suited for assessing variant prioritisation tools that will have a general applicability. Currently, the diagnostic rate for large-scale genomic RD studies is around 25% but this measure differs substantially depending on the patient phenotype, ranging from as low as 3% in patients with complex aetiologies to almost 50% in patients with neurodevelopmental disorders (21). However, the take home message is clearly that the majority of patients are left in a diagnostic odyssey and more work is needed to improve the diagnostic rate.

The purpose of this study is two-fold; firstly we want to test if the addition of CCR data and Dn status into current variant prioritisation pathways can improve the identification of pathogenic variants (thereby reducing time-taken-to-diagnosis) and secondly, we want to test whether this extra information helps to identify novel pathogenic variants.

## Methods

### Data access

The GeL research environment (RE) was used for all data analyses. We used the genomic data corresponding to data release v12, accessed through LabKey and extracted using RStudio(v1.4.1103) (22) library RLabkey. We used two sources of patient data for this project. First, we used the *gmc_exit_questionnaire* table which contains the diagnostic results provided by the clinical scientists from the Genomic Medicine Centres (GMCs) for each patient. This data informs to what extent a family’s presented case can be explained by the combined variants reported to the GMC from GeL and the Clinical Interpretation Providers. It also includes information on any segregation testing performed, the confidence in the identification and pathogenicity of each variant and, the clinical validity of each variant or variant pair in general and clinical utility in a specific patient. One Exit Questionnaire is completed per case.

Secondly, for the Dn variant analysis we used tables denovo_cohort_information which provided the ID information for each participant run through the Dn pipeline and denovo_flagged_variants which gives a list of all the Dn variants called and their confidence level. Full information on the GeL Dn variant research dataset is available at https://cnfl.extge.co.uk/display/GERE/De+novo+variant+research+dataset.

The phenotype data associated with the 100KGP participants is based on the Human Phenotype Ontology (HPO) terms (23) provided by the submitting GMC and is made available in the disease_phenotype table at three successive levels of detail; *Disease Group* gives a higher order description of the general phenotype class e.g. Skeletal disorders or Cardiovascular disorders, *Disease Sub Group* gives a finer scale description of the phenotype e.g. Skeletal dysplasias or Cardiomyopathy and, *Specific Disease* provides a detailed description of the specific disease e.g. Osteogenesis imperfecta or Dilated Cardiomyopathy.

For the Constrained Coding Region (9) analysis we downloaded the data from the website of Aaron Quinlan (https://s3.us-east-2.amazonaws.com/ccrs/ccrs/ccrs.autosomes.v2.20180420.bed.gz).

### Data cleaning

This was performed primarily using RStudio with the library tidyverse but also included substantial manual manipulation to harmonise the data to a consistent format. For the *gmc_exit_questionnaire*, patients were only retained if there was information for a genomic variant and the reference genome build. We classified patients in the ‘case_solved_family’ column as: Diagnosed (referred to as yes or partially), as each variant has been determined to be pathogenic by a clinical scientist or as Undiagnosed (referred to as no or unknown).

To build our cohort for analysis we split the data from the *gmc_exit_questionnaire* table into genome builds GRCh37 and GRCh38 and for both, we removed any duplicates and those variants from the X chromosome or mitochondrial genome. For the diagnosed patients with variants classed as Solved or Partial, we manually curated the data using the information available to leave only those variants that were pathogenic. For the undiagnosed patients with variants classed as no or Unknown, we kept all the variants that had been returned by the GMC. To unite the two genome builds into one cohort (gmc_ALL38), we used the UCSC Genome Browser LiftOver tool (http://genome.ucsc.edu/cgi-bin/hgLiftOver) and converted the coordinates for all GRCh37 variants into GRCh38.

To build our Dn cohort we first extracted the denovo_cohort_information table and only kept samples labelled as, member = Offspring and affection_status = AFFECTED. For the denovo_flagged_variants table we only kept variants with a Stringent Filter score =1. We then amended the denovo_flagged_variants table with participant_id information from the denovo_cohort table using the Trio Id as a scaffold and then split these samples by genome build GRCh37 and GRCh38. Using the UCSC Genome Browser LiftOver tool (http://genome.ucsc.edu/cgi-bin/hgLiftOver) we converted the GRCh37 variants into GRCh38 and combined the variants (Dn_ALL38).

As only a subset of the gmc_ALL38 were included in the Dn analysis, we used the participant_id information from DN_ALL38 to derive a final cohort that contained diagnostic information from only those patients present in both cohorts, this is the cohort we used for our analysis (gmc_Dn_ALL).

To identify which of the variants from the gmc_Dn_ALL cohort were Dn we used the Dn_ALL38 data and extracted those variants that matched for participant_id and genomic position (Dn_gmc).

For the CCR intersection analysis we first removed all the VARTRUE variants from the bed file as these correspond to known variants in the gnomAD database. Because the CCR region coordinates are in genome build GRCh37 we used the UCSC Genome Browser LiftOver tool to convert the coordinates to GRCh38. We then used bedtools (24) intersect to extract those variants that resided within a CCR and included their percentile score in the output. Using the percentile score we were able to filter our variants to only those that intersected a CCR with a percentile score of ≥95 (CCR95).

Combining the results of the Dn_gmc and CCR95 intersection we were able to derive a list of Dn variants from the gmc_ALL38 cohort that intersected a CCR95 (Dn_CCR95). Specifically for this annotation category, we performed a re-evaluation of the Undiagnosed patients within a research context. We first extracted the patient’s phenotype information from the GeL RE using the rare_disease_participant_phenotype table and then, for each variant we annotated it using wANNOVAR (25) before performing a literature search using PubMed (https://pubmed.ncbi.nlm.nih.gov/), OMIM (https://omim.org/) and ClinVar (11), to evaluate the evidence linking the variant to the patient’s phenotype. Where the evidence suggested the variant was linked to the phenotype and a diagnosis was probable, we submitted the information to GeL who contacted the referring clinician or GMC for a formal clinical evaluation.

### Disease terms

We used the patient_phenotype table in LabKey to extract the phenotype information for the gmc_ALL38 cohort. The disease terms are provided at three levels starting broad and becoming more focused (*Disease Group, Disease Sub Group and Specific Disease*). As the disease term assigned to each patient becomes more specific the overall number of terms increases greatly and consequently the number of patients within each term decreases. We therefore set a cut-off ≥5 patients per term and chose to focus on the highest descriptive level, *Disease Group* to ensure we had sufficient numbers in each group to make meaningful comparisons.

### Analysis strategy

We focused on assessing the proportion of variants present in the following three annotation categories; 1) Dn variant, 2) intersecting a CCR95 and, 3) a Dn variant intersecting a CCR95. The aims of this were twofold: Firstly we wanted to explore whether there was an enrichment of pathogenic variants in one of the three annotation categories to examine whether they could be used as an additional filter to identify pathogenic variants more specifically and thus potentially allow a diagnosis to be reached quicker. Secondly, we wanted to explore if, in the undiagnosed samples, the combined application of the Dn and CCR95 annotations would filter the number of variants down to a small enough number to highlight novel disease variants.

Furthermore, we also explored whether the analysis strategy above was applicable to RDs across the clinical spectrum or whether certain disease phenotypes were more applicable to this approach by splitting our cohort into separate disease classes and looking for enrichment of signal. To do this we first calculated the number of diagnosed and undiagnosed patients in the total cohort (including those in Disease Groups with <5 patients). Our null hypothesis was that the same proportion of diagnosed and undiagnosed patients should be represented in the three annotation categories (Dn, CCR95 and Dn/CCR95). For each annotation category we calculated the number of Diagnosed patients we expect for each Disease Group and compared it to the observed number using a Chi-square test to derive a p-value. We then used a Bonferroni correction to account for multiple testing (10 x Disease Groups and 3 x annotation categories = 30 tests) to derive an adjusted p-value (p_adj).

## Results

### Sample cohort

For our analyses we used the 100KGP v12 dataset which is comprised of 73,880 RD genomes from 71,597 participants. Following mapping and variant calling (21), 33,315 families have been processed through an automated analytic pipeline to filter down the variants to rare, segregating and predicted damaging candidate variants in coding regions. These variants have been classed as tier 1, 2 or 3 (see (21) for full description) and used to make a clinical diagnosis in 30,419 families.

Although Dn status can be predicted through simple analysis of the trio vcf file data it does not provide a robust output. Therefore, for our Dn analyses we analysed 13,353 trios using a bespoke Dn pipeline that utilised the raw sequence data for each member of the trio.

The cohort we used for this analysis was derived from the overlapping patients that had undergone a clinical diagnosis and had been run through the bespoke Dn analysis pipeline (n = 3,631). However, because the CCR data has been generated for the autosomes only, we removed all samples with a variant assigned to the X-Chromosome or mitochondrial genome which resulted in a final cohort of 3,090 families.

### Diagnostic rate calculation

Following data cleaning we calculated the diagnostic rate for the gm_Dn_ALL cohort as a whole was 71% (Table 1). When we stratify the data for Dn CCR95 annotation, we show the causative variant is Dn in 40% of patients, intersects with a CCR95 in 18% of the patients and is a Dn intersecting a CCR95 in 12% of patients (Table 1).

**Table 1.**
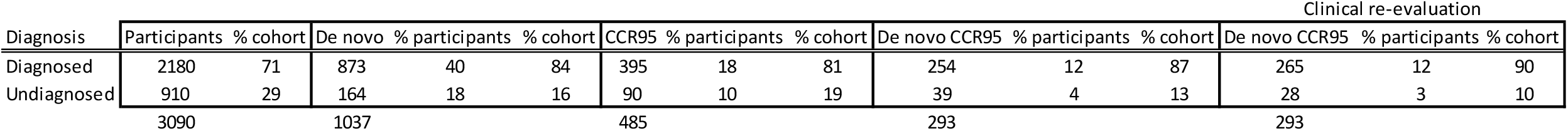
Table showing the numbers and percentage of diagnosed and undiagnosed patients in the full cohort for each annotation class. The Clinical re-evaluation column provides the data following a clinical re-evaluation of the variants from the undiagnosed patients from within the Dn CCR95 annotation category with strong evidence for being disease causing.

If we next look at each annotation class and calculate the diagnostic rate within each group, we see that the diagnostic rate increases to 84% for Dn variants, 81% for CCR95 variants and 87% for Dn CCR95 intersecting variants (Table 1). That is, for example, if we extract all the Dn CCR95 intersecting variants (n=293), 254 of these (87%) have been classed as pathogenic by a clinical scientist, which is a highly significant enrichment p=6.42×10^-6^ (Supplementary Table 1).

We next focused in on the undiagnosed patients who had a Dn CCR95 intersecting variant, to search for novel diagnoses (n=39). Our research-based analysis identified 27 variants with strong evidence supporting them as potential diagnostic variants based on the patients phenotype and the genomic function of the variant. Thus far, we have had clinical confirmation that 11 of these variants (41%) have been classed as causative, which increases the proportion of Dn CCR95 variants that are diagnostic to 90%.

Further stratification of our data by disease terms revealed that the patients with a phenotype within the Neurology and neurodevelopmental disorders domain showed a highly significant enrichment of pathogenic variants in all annotation classes (Table 2). This ranged from 53% in the whole cohort to 69% in the Dn cohort (Enrichment p_adj = 2.24×10^-10^), 76% in CCR95 cohort (Enrichment p_adj = 7.47×10^-10^) and 80% in the Dn_CCR95 cohort (Enrichment p_adj = 9.48×10^-09^).

**Table 2.** Table showing the enrichment of specific Disease Groups for each annotation category compared to the full cohort. The p_adj column shows the significance values for only those Disease Groups with an adjusted p-value ≤0.05. * This value reflects the total number of participants in the full cohort, those Disease Groups with <5 participants are not included in this table.

| Disease Group | All |  | % of total |  | De novo |  |  |  | CCR95 |  |  |  | De novo CCR95 |  |  |  |
| --- | --- | --- | --- | --- | --- | --- | --- | --- | --- | --- | --- | --- | --- | --- | --- | --- |
|  | Diagnosed | Undiagnosed | Diagnosed | Undiagnosed | exp | obs | p-value | p_adj | exp | obs | p-value | p_adj | exp | obs | p-value | p_adj |
| Endocrine disorders | 55 | 25 | 2 | 3 | 22 | 16 | 0.33 |  | 10 | 7 | 0.46 |  | 6 | 4 | 0.52 |  |
| Dysmorphic and congenital abnormality syndromes | 56 | 29 | 3 | 3 | 22 | 29 | 0.32 |  | 10 | 14 | 0.41 |  | 6 | 8 | 0.59 |  |
| Cardiovascular disorders | 59 | 61 | 3 | 7 | 23 | 22 | 0.88 |  | 11 | 7 | 0.34 |  | 7 | 4 | 0.36 |  |
| Metabolic disorders | 66 | 21 | 3 | 2 | 26 | 22 | 0.56 |  | 12 | 6 | 0.15 |  | 8 | 5 | 0.4 |  |
| Renal and urinary tract disorders | 80 | 39 | 4 | 4 | 32 | 14 | 0.0072 |  | 14 | 4 | 0.017 |  | 9 | 0 | 0.0025 |  |
| Hearing and ear disorders | 81 | 50 | 4 | 5 | 32 | 13 | 0.0041 |  | 15 | 6 | 0.047 |  | 9 | 3 | 0.08 |  |
| Skeletal disorders | 120 | 26 | 5 | 3 | 48 | 41 | 0.45 |  | 22 | 5 | 0.00087 | 0.026 | 14 | 2 | 0.0023 |  |
| Ultra-rare disorders | 140 | 74 | 6 | 8 | 56 | 61 | 0.63 |  | 25 | 24 | 0.88 |  | 16 | 16 | 1 |  |
| Ophthalmological disorders | 227 | 49 | 10 | 5 | 90 | 22 | 3.13E-11 | 9.39E-10 | 41 | 7 | 4.13E-07 | 1.24E-05 | 26 | 3 | 1.09E-05 | 0.00033 |
| Neurology and neurodevelopmental disorders | 1177 | 510 | 53 | 54 | 467 | 607 | 7.45E-12 | 2.24E-10 | 211 | 301 | 2.49E-11 | 7.47E-10 | 135 | 202 | 3.16E-10 | 9.48E-09 |
|  | 2214* | 938* |  |  |  | 879* |  |  |  | 397* |  |  |  | 254* |  |  |

In comparison, those patients with a phenotype in the Ophthalmological disorders domain (the next largest *Disease Group*), showed a highly significant negative enrichment in all annotation classes (Table 2). This group comprised 10% of the whole cohort, 3% of the Dn cohort (Enrichment p_adj = 9.39×10^-10^), 2% in the CCR95 cohort (Enrichment p_adj = 1.24×10^-05^) and 1% of the Dn_CCR95 cohort (Enrichment p_adj = 3.33×10^-04^) (see Supplementary Table 2 for all enrichment p-values).

## Discussion

Our study shows that selecting genetic variants with a Constrained Coding Region percentile score ≥95 (CCR95) in the variant prioritisation cascade used to clinically diagnose RD patients enriches for pathogenic variants (from 71% to 81%). This effect is similar to the enrichment seen when taking Dn status into account (from 71% to 84%) and is further increased when combing both metrics, that is, Dn variants residing within a CCR95 (from 71% to 87%) (Table 1). We hypothesise that incorporating this metric into the genomic clinical diagnostic filtering cascade would decrease the time taken to identify pathogenic variants and therefore allow more patients to be screened over a set period of time, thus resulting in an increase in the diagnostic rate.

A major advantage of the CCR method is that it offers a focused annotation metric for discrete regions of a gene instead of methods such as pLI (probability of being loss-of-function intolerant) (26) or Z scores (27) that are provided in gnomAD (10), which annotate the whole gene. The reason this is important is because, even in highly constrained genes (pLI ≥ 0.9, positive Z score), there are often discrete regions that show low constraint with many functional variants seen in gnomAD (10). Therefore, if a potentially pathogenic variant is located in one of these low constrained regions the variant could be misinterpreted as pathogenic based on the gene-wide annotation. Conversely, there are instances where genes showing low constraint (pLI ≤ 0.5, negative Z score) contain small, highly constrained regions that are devoid of functional variation in gnomAD (10) which can lead to a pathogenic variant, from within this region, being missed if the gene-wide annotation is used (see (9) Figure 1).

Furthermore, because the number of variants fulfilling the Dn_CCR95 criteria is low, it is feasible to use this metric to identify novel pathogenic variants. For example, we identified 39 variants in patients classed as Undiagnosed that were in the Dn_CCR95 category (Table 1). Inspection of these variants in a research setting led to a candidate diagnosis for 27 of the undiagnosed patients (69%) (Supplementary Table 3).

For example, in an undiagnosed patient with HPO (23) terms including microcephaly, intellectual disability, small for gestational age, short stature, motor axonal neuropathy and congenital microcephaly; we identified a missense variant in the gene *MORC2* (p.Y448C) that was predicted to be deleterious by SIFT (28), probably damaging by PolyPhen-2 (29) and had a CADD (30) Phred score of 30. This variant is also predicted to be likely pathogenic in ClinVar (Variation ID: 804228). Patients with *MORC2* pathogenic variants present with a phenotype consisting of developmental delay, intellectual disability, growth retardation, microcephaly, variable craniofacial dysmorphism, and in some individuals electrophysiologic abnormalities suggestive of neuropathy (31). The phenotypic overlap along with the damaging nature of this variant make this a strong candidate to be the pathogenic variant for this patient.

The information for all 27 candidate diagnostic variants was fed back via GeL to the clinicians who submitted these patients for a clinical re-evaluation. Thus far, this approach has resulted in an extra 11 patients receiving a clinical diagnosis (including *MORC2*) which improves the diagnostic rate for De novo CCR95 variants to 90% (Table 1). Our analysis also provides compelling evidence that a further 8 variants are diagnostic but we currently lack a clinical interpretation of these (supplementary table 3). We therefore believe, the proportion of Dn CCR95 variants that are diagnostic is likely to be in excess of 90% in this cohort.

An example of this includes an undiagnosed patient from the Renal and urinary tract disorders Disease Group that had a missense variant in the gene *SETD5* (p.R348L) which was predicted to be deleterious by SIFT (28), probably damaging by PolyPhen-2 (29) and had a CADD (30) Phred score of 32. This variant is also predicted to be likely pathogenic in ClinVar (Variation ID: 1065941). The HPO (23) terms for this patient included abnormality of the eye, hypodontia, pectus excavatum, global developmental delay, abnormal facial shape and mild short stature, none of which fit with the Disease Group they were assigned to. Patients with *SETD5* pathogenic variants display variable features including intellectual disability, facial dysmorphism, cardiac and skeletal abnormalities, behavioural problems and short stature (32). We therefore believe the *SETD5* variant in this patient is likely to be pathogenic.

Although we cannot be sure why this patient was included in the Renal and urinary tract disorders Disease Group, this example highlights the power of combining Dn and CCR95 data as it allows a disease/gene agnostic approach to identify highly likely pathogenic variants. This approach will therefore help in overcoming problems of misdiagnoses or when patients have multi-system abnormalities and do not fit a single phenotype grouping. Also, because applying this filter results in very few variants qualifying for inspection, it’s use as a first-pass stringent filter in clinical diagnostic laboratories has the potential to rapidly identify pathogenic variants from patient cohorts, freeing up time to screen more patients overall.

To explore our data further we next performed an *ad hoc* analysis to search recurrent Dn sites for instances where a Dn had been called pathogenic in one patient but not in another or where a recurrent Dn was seen in greater than one undiagnosed patient (Supplementary Table 4). We reasoned that this approach could help diagnose patients where, for whatever reason, there was not enough evidence to call a variant pathogenic. We identified 44 Dn variants that were recurrent in 100 individuals. Of these, 36 were identified in only diagnosed patients, six were a mixture where at least one patient was diagnosed and the other was undiagnosed, and in two instances, where both patients were undiagnosed.

For the latter group, our analysis highlighted a Dn canonical splice site variant (c.2493+1 G>A) in the Floating-Harbour syndrome gene (33) *SRCAP* (NM_006662.3) in two undiagnosed patients with intellectual disability. The evidence available (CADD Phred score 36) makes this a highly likely pathogenic variant in these patients. At the second loci, we identified two undiagnosed patients with a Dn missense variant in the gene *FBXO11* in which Dn variants are known to cause an intellectual developmental disorder with dysmorphic facies and behavioural abnormalities (IDDFBA) (34). The Dn variant we identified at codon 54 (NM_025133) causes a change from an Arginine to Glycine amino acid which is not present in gnomAD (10), is predicted by SIFT (28) to be tolerated, benign by PolyPhen-2 (29) and has a CADD (30) Phred score of 20. In ClinVar there is a known Pathogenic/Likely pathogenic missense variant (ClinVar ID:559601) at this codon which causes an amino acid change from Arginine to Serine which is also not present in gnomAD (10). Arginine is a basic amino acid and therefore a change to Glycine (small) or Serine (nucleophilic) could both potentially have functional effects, especially as they occur within a disorganised protein domain. However, the predicted deleteriousness of the Dn variant we identified is weak and the CCR percentile score for this region is 68, meaning we would not be confident in calling this variant pathogenic/likely pathogenic without additional evidence.

Of the recurrent Dn variants where one patient remains undiagnosed, our analysis provides evidence for a diagnosis in a further 4 patients (supplementary table 4). This includes an undiagnosed patient with a variant in the gene *CHD4*. This patient’s variant was also included in the Dn CCR95 annotation category and following clinical re-evaluation was deemed to be diagnostic (supplementary table 3). This demonstrates the effectiveness of this approach and if replicated for the other recurrent Dn variants also highlighted as having strong evidence for pathogenicity, would provide a diagnosis for at least another 5 patients.

Although our study shows the benefit of incorporating CCR percentile score information and Dn status in the filtering cascade for identifying pathogenic RD variants we need to be aware of the constraints relating to the cohort we have used. Firstly, in the cohort we have studied the diagnostic rate was far higher (71%) than that seen in other RD cohorts, including the 100KGP pilot project (25%) (21). This may be due to many reasons; first, the data in the 100KGP gmc_exit_questionnaire table contains entries for 30,419 families, however, many of these entries have no genomic annotations, most of which are from the unsolved group (21,048). This means that once we had filtered the data to include only patients that were also included in the Dn cohort and filtered out any variants on the X chromosome and mitochondrial genome we had a cohort of 3,090 patients for our analysis. Because the variants contained in the gmc_exit_questionnaire are returned by the gmc centres that recruited the patients following diagnostic evaluation, we can assume that no Tier 1 or Tier 2 variants were present in the undiagnosed patients that would explain their phenotype. The Tiering of the data is performed by GeL commercial partners and so without reanalysing the whole dataset from the raw data we are unable to estimate how many Dn or CCR95 variants may be present in these un-annotated patients.

Secondly, the patients recruited to the 100KGP were partly composed of research cohorts and patients who previously had negative genetic tests and therefore may represent a distinct cohort of patients not fully representative of the diverse patients seen in NHS clinical diagnostic centres. To truly estimate the clinical utility of applying Dn and CCR95 status we would need to incorporate these annotations into a routine NHS diagnostic laboratory patient cohort and perform a direct comparison of the diagnostic rate with these annotations and without them. This would also allow us to estimate if the application of these annotations decreases the time taken to reach a diagnosis and if so how much it improves the overall diagnostic rate. If this was to be done, it would be imperative we include CCR annotations for the X chromosome as we could potentially be missing out on a number of diagnoses that reside on that chromosome.

Finally, the inclusion of Disease Group data showed there was a large bias towards patients with a neurology and neurodevelopmental disorders phenotype (>50% of the cohort) which may have biased the results somewhat (Table 2). This is especially pertinent for the Dn analysis as Dn variants are a well-known source of pathogenicity for patients with such phenotypes (12-18), as shown by the highly significant enrichment we saw of Dn variants in this group (p_adj = 2.24×10^-10^). It should be noted however, that we also observed a highly significant enrichment of variants from this group that intersected a CCR95 region (p_adj = 7.47×10^-10^), 28% of which were not Dn variants.

This observation is in stark contrast to the pattern of enrichment we saw for patients within the ophthalmological disorders Disease Group. For these patients we saw a highly significant underrepresentation of Dn, CCR95 and Dn CCR95 variants (Table 2). For patients within the skeletal disorders Disease Group, we observed an underrepresentation of variants intersecting CCR95 regions which was also observed for Dn CCR95 variants but not for Dn variants alone (Table 2). For the remaining Disease Group phenotypes, the numbers were much smaller which reduced our power to identify any significant enrichments.

Why we see such an enrichment for Dn and CCR95 variants in the neurology and neurodevelopmental disorders Disease Group patients is open to speculation but is beyond the scope of this study. Nonetheless, it does suggest that for patients with a neurology and neurodevelopmental disorders phenotype, the greatest diagnostic return will come from a trio sequencing approach and that this should be the first-tier test used by clinical diagnostic laboratories.

Where a Dn analysis is not possible, due to unavailability of one or both parents or because of cost constraints, it is noteworthy that the use of CCR95 status alone will enable the identification of highly likely pathogenic variants. Our analysis shows that, of all the variants that intersected a CCR95 (n=485), 81% (n=395) were clinically diagnosed pathogenic variants (Table 1). This observation could be particularly relevant to clinical diagnostic laboratories in low/middle economic countries where sequencer availability and costs may render a Dn-trio diagnostic approach unfeasible for patients with a neurology and neurodevelopmental disorders phenotype.

In summary, we have constructed a cohort of RD patients (n=3,090) that have undergone WGS as part of the GeL 100KGP, have been clinically assessed by a diagnostic laboratory and who have undergone a bespoke Dn variant calling pipeline. We have sought to determine if taking into account a variant’s Dn status, whether it intersects a CCR95 or both could improve our ability to identify clinically pathogenic variants. We show how the rate of diagnosis in our whole cohort (71%) increases when we look at just those variants that are classed as intersecting a CCR95 (81%), Dn variants (84%) or a combination of both (90%) (Table 1). This observation seems to be driven primarily by patients with a neurology and neurodevelopmental disorders phenotype. We therefore suggest that, where possible patients with such a phenotype should receive trio WGS as a first tier test but where parent availability or cost make this unfeasible, the identification of variants intersecting a CCR95 is an alternative route to aid in the detection of highly likely clinically pathogenic variants.

Further work should aim to apply this approach in systematic way in a routine clinical diagnostic laboratory to assess its utility and to determine if it can decrease the time taken to reach a diagnosis and therefore, allow more diagnoses to be made and thus increase the rate of diagnoses overall.

## Data Availability

All data produced in the present work are contained in the manuscript

## Acknowledgements

This research was made possible through access to the data and findings generated by the 100,000 Genomes Project. The 100,000 Genomes Project is managed by Genomics England Limited (a wholly owned company of the Department of Health). The 100,000 Genomes Project is funded by the National Institute for Health Research and NHS England. The Wellcome Trust, Cancer Research UK and the Medical Research Council have also funded research infrastructure. The 100,000 Genomes Project uses data provided by patients and collected by the National Health Service as part of their care and support.

---

**Supplementary Table 1.** Table showing the values used to derive the enrichment of diagnosed patients in each annotation class compared to the whole cohort.

| Diagnosis | Participants | % cohort | expected | observed | De novo |  |
| --- | --- | --- | --- | --- | --- | --- |
|  |  |  |  |  | chi-square p-value | Bonferroni corrected p-value (x3 tests) |
| Diagnosed | 2180 | 71 | 732 | 873 | 1.35E-13 | 4.05E-13 |
| Undiagnosed | 910 | 29 | 305 | 164 |  |  |
| Total | 3090 |  |  | 1037 |  |  |

| Diagnosis | Participants | % cohort | expected | observed | CCR95 |  |
| --- | --- | --- | --- | --- | --- | --- |
|  |  |  |  |  | chi-square p-value | Bonferroni corrected p-value (x3 tests) |
| Diagnosed | 2180 | 71 | 342 | 395 | 6.79E-05 | 2.04E-04 |
| Undiagnosed | 910 | 29 | 143 | 90 |  |  |
| Total | 3090 |  |  | 485 |  |  |

| Diagnosis | Participants | % cohort | expected | observed | De novo CCR95 |  |
| --- | --- | --- | --- | --- | --- | --- |
|  |  |  |  |  | chi-square p-value | Bonferroni corrected p-value (x3 tests) |
| Diagnosed | 2180 | 71 | 207 | 254 | 2.14E-06 | 6.42E-06 |
| Undiagnosed | 910 | 29 | 86 | 39 |  |  |
| Total | 3090 |  |  | 293 |  |  |

| Diagnosis | Participants | % cohort | expected | observed | De novo CCR95* |  |
| --- | --- | --- | --- | --- | --- | --- |
|  |  |  |  |  | chi-square p-value | Bonferroni corrected p-value (x3 tests) |
| Diagnosed | 2180 | 71 | 207 | 264 | 7.79E-07 | 2.34E-06 |
| Undiagnosed | 910 | 29 | 86 | 29 |  |  |
| Total | 3090 |  |  | 293 |  |  |

\* Updated with diagnostic results for undiagnosed Dn CCR patients

**Supplementary Table 2.**
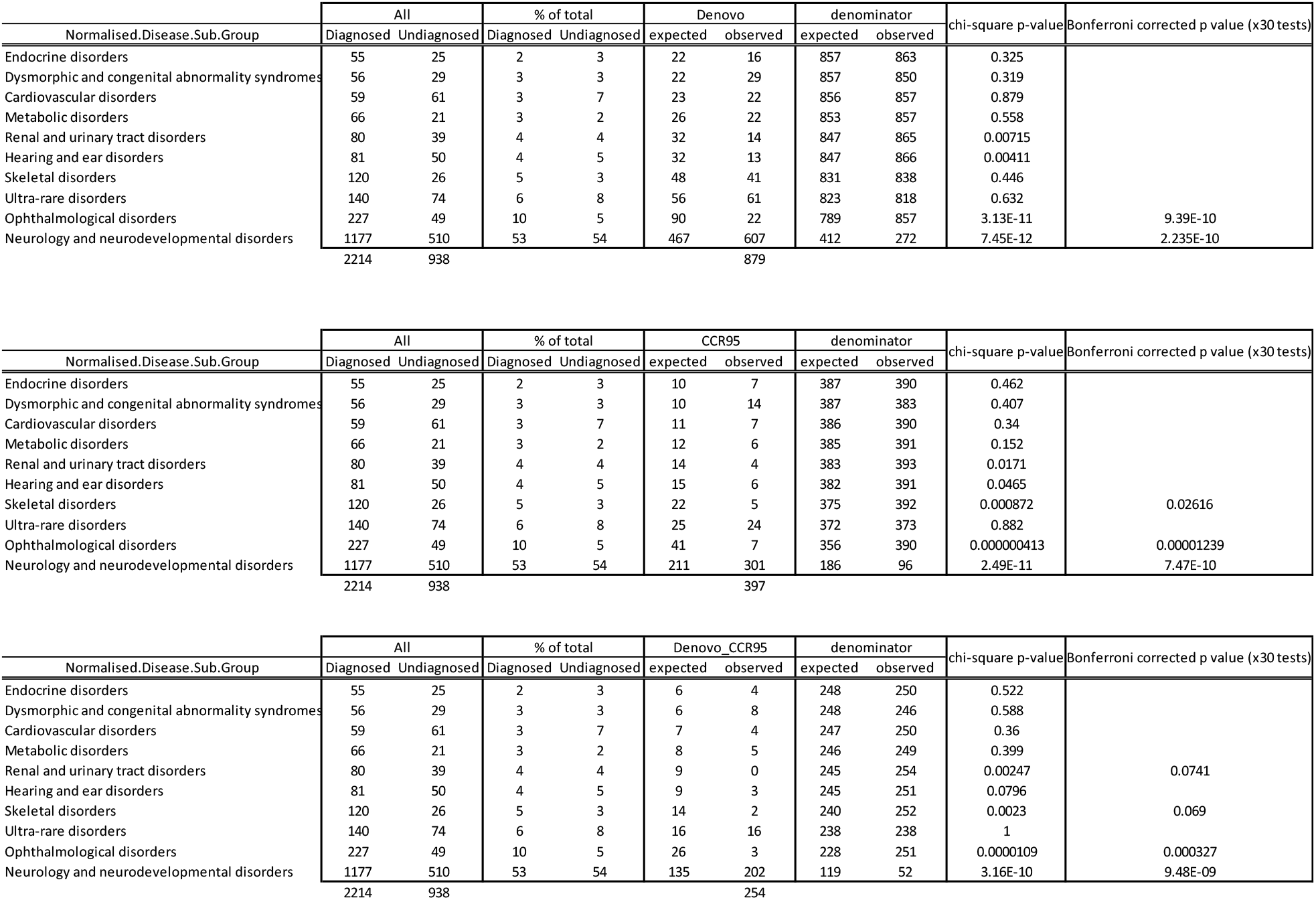
Table showing the values used to derive the enrichment of Disease Groups within each of the annotation classes compared to the whole cohort.

**Supplementary Table 3.** Table showing the 39 variants from the undiagnosed patients within the Dn CCR annotation category. Following a research-based evaluation the variants were classed according to whether the variant was a good phenotypic fit for the patient based on current literature (see highlighted references). Those that did fit the phenotype were returned to Genomics England for a clinical re-evaluation, the ‘Diagnostic’ variants confirm the variant has been clinically assessed as causative for the patient and ‘Strong evidence’ refers to variants that have compelling evidence but are awaiting a clinical re-evaluation.

| Chromosome | Position | Reference | Alternate | Gene Name | CCR Percentile | Disease Group | Phenotype fit | Reference | Re-evaluation |
| --- | --- | --- | --- | --- | --- | --- | --- | --- | --- |
| 1 | 244864247 | T | C | HNRNPU | 96 | Neurology and neurodevelopmental disorders | Yes | PMID: 28393272 | Strong evidence |
| 2 | 165344802 | G | A | SCN2A | 99 | Neurology and neurodevelopmental disorders | Yes | PMID: 33985586 | Diagnostic |
| 2 | 165381126 | C | A | SCN2A | 99 | Neurology and neurodevelopmental disorders | Yes | PMID: 33985586 |  |
| 2 | 165389042 | T | C | SCN2A | 97 | Neurology and neurodevelopmental disorders | Yes | PMID: 32929885 |  |
| 2 | 165389115 | T | A | SCN2A | 99 | Neurology and neurodevelopmental disorders | Yes | PMID: 32929885 | Diagnostic |
| 3 | 9442211 | G | T | SETD5 | 99 | Renal and urinary tract disorders | Yes | PMID: 34169511 | Strong evidence |
| 3 | 11025521 | G | A | SLC6A1 | 97 | Neurology and neurodevelopmental disorders | Yes | PMID: 34006619 | Diagnostic |
| 3 | 53660224 | C | T | CACNA1D | 98 | Neurology and neurodevelopmental disorders | Yes | PMID: 33985586 |  |
| 4 | 73102546 | A | C | ANKRD17 | 100 | Neurology and neurodevelopmental disorders | Yes | PMID: 33909992 | Diagnostic |
| 4 | 73120891 | T | C | ANKRD17 | 99 | Neurology and neurodevelopmental disorders | Yes | PMID: 33909992 | Diagnostic |
| 6 | 3227539 | C | A | TUBB2B | 99 | Neurology and neurodevelopmental disorders | Yes | PMID: 32570172 | Diagnostic |
| 6 | 90556536 | C | G | MAP3K7 | 97 | Ultra-rare disorders | Yes | PMID: 34558790 |  |
| 7 | 98955183 | C | T | TRRAP | 98 | Neurology and neurodevelopmental disorders | Yes | PMID: 30827496 | Diagnostic |
| 9 | 83970194 | C | G | HNRNPK | 99 | Neurology and neurodevelopmental disorders | Yes | PMID: 32588992 | Strong evidence |
| 9 | 83977032 | C | T | HNRNPK | 99 | Dysmorphic and congenital abnormality syndromes | Yes | PMID: 32588992 | Strong evidence |
| 9 | 127678531 | A | AG | STXBP1 | 99 | Neurology and neurodevelopmental disorders | Yes | PMID: 34392114 | Strong evidence |
| 10 | 252399 | T | C | ZMYND11 | 99 | Neurology and neurodevelopmental disorders | Yes | PMID: 32097528 |  |
| 12 | 6587934 | G | A | CHD4 | 100 | Neurology and neurodevelopmental disorders | Yes | PMID: 24991957 | Diagnostic |
| 14 | 102008244 | C | T | DYNC1H1 | 97 | Neurology and neurodevelopmental disorders | Yes | PMID: 38513047 | Strong evidence |
| 16 | 8906511 | T | C | USP7 | 98 | Neurology and neurodevelopmental disorders | Yes | PMID: 30679821 |  |
| 17 | 8535898 | C | T | MYH10 | 97 | Neurology and neurodevelopmental disorders | Yes | PMID: 30879067 | Strong evidence |
| 17 | 29480472 | C | T | TAOK1 | 100 | Neurology and neurodevelopmental disorders | Yes | PMID: 33565190 |  |
| 19 | 11025448 | G | A | SMARCA4 | 100 | Neurology and neurodevelopmental disorders | Yes | PMID: 34180430 |  |
| 19 | 12865105 | G | A | MAST1 | 99 | Neurology and neurodevelopmental disorders | Yes | PMID: 31721002 | Diagnostic |
| 19 | 13262778 | C | T | CACNA1A | 99 | Neurology and neurodevelopmental disorders | Yes | PMID: 14718690 | Diagnostic |
| 20 | 63495939 | C | T | EEF1A2 | 100 | Neurology and neurodevelopmental disorders | Yes | PMID: 32196822 |  |
| 22 | 20991687 | G | T | LZTR1 | 96 | Dysmorphic and congenital abnormality syndromes | Yes | PMID: 34184824 | Strong evidence |
| 22 | 30958692 | G | A | MORC2 | 96 | Ultra-rare disorders | Yes | PMID: 32693025 | Diagnostic |
| 1 | 244856619 | C | T | HNRNPU | 95 | Ultra-rare disorders | No |  |  |
| 3 | 24122913 | G | C | THRB | 99 | Neurology and neurodevelopmental disorders | No |  |  |
| 3 | 25569883 | T | G | RARB | 96 | Ophthalmological disorders | No |  |  |
| 8 | 42472309 | C | T | SLC20A2 | 96 | Neurology and neurodevelopmental disorders | No |  |  |
| 8 | 116854385 | G | A | RAD21 | 98 | Dysmorphic and congenital abnormality syndromes | No |  |  |
| 10 | 110581019 | C | T | SMC3 | 99 | Ultra-rare disorders | No |  |  |
| 12 | 110327992 | C | A | ATP2A2 | 99 | Cardiovascular disorders | No |  |  |
| 12 | 70346226 | AC | A | CNOT2 | 100 | Ophthalmological disorders | No |  |  |
| 15 | 40728770 | C | T | RAD51 | 95 | Neurology and neurodevelopmental disorders | No |  |  |
| 20 | 58909375 | C | T | GNAS | 98 | Neurology and neurodevelopmental disorders | No |  |  |
| 21 | 33568984 | C | T | SON | 96 | tumour syndromes | No |  |  |

**Supplementary Table 4.** Table showing the recurrent Dn variants and whether following a research-based evaluation there is sufficient evidence to support a potential diagnosis. * This variant has been classed as diagnostic following a clinical re-evaluation as it is also part of the Dn CCR annotation category (see supplementary table 3).

| Chromosome | Position | Reference | Alternate | Gene name | ACMG classification | Disease group | Diagnosis status | Potential diagnosis |
| --- | --- | --- | --- | --- | --- | --- | --- | --- |
| 7 | 40047840 | G | C | CDK13 | likely_pathogenic_variant | Neurology and neurodevelopmental disorders | Diagnosed |  |
| 7 | 40047840 | G | C | CDK13 | likely_pathogenic_variant | Skeletal disorders | Undiagnosed | No |
| 11 | 687941 | C | T | DEAF1 | pathogenic_variant | Neurology and neurodevelopmental disorders | Diagnosed |  |
| 11 | 687941 | C | T | DEAF1 | likely_pathogenic_variant | Neurology and neurodevelopmental disorders | Undiagnosed | Yes |
| 12 | 6587934 | G | A | CHD4 | likely_pathogenic_variant | Neurology and neurodevelopmental disorders | Diagnosed |  |
| 12 | 6587934 | G | A | CHD4 | likely_pathogenic_variant | Neurology and neurodevelopmental disorders | Undiagnosed | Yes* |
| 12 | 112477720 | A | G | PTPN11 | pathogenic_variant | Hearing and ear disorders | Diagnosed |  |
| 12 | 112477720 | A | G | PTPN11 | pathogenic_variant | Neurology and neurodevelopmental disorders | Diagnosed |  |
| 12 | 112477720 | A | G | PTPN11 | pathogenic_variant | Hearing and ear disorders | Undiagnosed | No |
| 17 | 76137651 | C | - | FOXJ1 | pathogenic_variant | Ciliopathies | Diagnosed |  |
| 17 | 76137651 | C | - | FOXJ1 | VUS | Ciliopathies | Undiagnosed | Yes |
| 20 | 32436955 | C | T | ASXL1 | likely_pathogenic_variant | Neurology and neurodevelopmental disorders | Diagnosed |  |
| 20 | 32436955 | C | T | ASXL1 | likely_pathogenic_variant | Neurology and neurodevelopmental disorders | Undiagnosed | Yes |
| 2 | 47839449 | T | C | FBXO11 | VUS | Neurology and neurodevelopmental disorders | Undiagnosed | No |
| 2 | 47839449 | T | C | FBXO11 | VUS | Neurology and neurodevelopmental disorders | Undiagnosed | No |
| 16 | 30713712 | G | A | SRCAP | VUS | Neurology and neurodevelopmental disorders | Undiagnosed | Yes |
| 16 | 30713712 | G | A | SRCAP | VUS | Neurology and neurodevelopmental disorders | Undiagnosed | Yes |

## Notes

### Competing Interest Statement

The authors have declared no competing interest.

### Funding Statement

This study did not receive any funding

### Author Declarations

Ethics committee of Genomics England plc gave ethical approval for this work

### Summary of Updates

This revision includes updated diagnostic rates following clinical re-evaluation of genomic variants from undiagnosed patients that were submitted to Genomics England and subsequently classed as diagnostic.

